# A Cross-Sectional Study on HPV Vaccine Awareness, Vaccination Willingness and Associated Factors among Male Healthcare Workers in Ethnic Minority-populated Areas in Southern China

**DOI:** 10.1101/2024.09.02.24312968

**Authors:** Chunlin Qin, Nian Jiang, Guorong Tang, Yun Zhou, Qingqing Liang

**Affiliations:** Guilin Center for Disease Control and Prevention, Guilin 541001, China; College of Public Health, Guilin Medical University, Guilin 541001,China; Guilin Health and Family Planning Information Centre, Guilin 541001,China

**Keywords:** Human Papilloma Virus, vaccination, awareness, willingness, associated factors, male healthcare workers, China

## Abstract

**Introduction:** Human papillomavirus (HPV) is a common sexually transmitted disease (STD) with a very high prevalence in the male population, resulting in an increasing burden of HPV-related diseases. HPV vaccines are the most effective measure to control HPV infection. However, male HPV vaccine has not been approved for mainland China.

**Objective:** To assess awareness, willingness and associated factors of the HPV vaccine among male healthcare workers in ethnic minority-populated areas in southern China. To provide a reference basis for the formulation of promotion strategy for male HPV vaccine after its approval in mainland China.

**Methods:** A web-based questionnaire survey on the awareness and willingness to receive HPV vaccine among male healthcare workers in the surveyed areas was conducted using a convenience sampling method, and chi-square test or Fisher’ s exact test and logistic regression analyses were used to analyze and explore the associated factors.

**Results:** Respondents’ HPV vaccine awareness and willingness to vaccinate were 74.55% (1,066/1,430) and 80.94% (1,087/1,343, exclusion of vaccinated respondents) respectively. Respondents aged 30∼44 and ≥ 45, intermediate title, and per capita monthly household income of CNY 3,000∼4,000 were discovered to have a correlation with awareness of HPV vaccine (all *p*<0.05). While, technicians and other occupations (excluding doctors and nurses), with a position, senior title, and ware of HPV vaccine were discovered to have a correlation with willingness to vaccinate (all *p*<0.05). The belief that the vaccine can prevent HPV infection was the main reason for participants’ willing to get HPV vaccine (91.48% 1,074/1,174). while, believing that they are not at risk of contracting HPV and the high price of HPV vaccine are major barriers to increasing willingness to vaccinate. The most important motivation that could promote receiving the vaccine was the provision of information on the efficacy and safety of the HPV vaccine.

**Conclusions:** Male healthcare workers have a high awareness of HPV vaccine and a high willingness of for HPV vaccination in the surveyed areas. Strengthening health education, including HPV vaccine in immunization programmes or health insurance subsidies, and lowing the cost of vaccination will help increase their willingness for HPV vaccination.

## 1. Introduction

Human papillomavirus (HPV) can infect human epithelial cells, including genital and oral mucosal epithelium, posing a serious threat to human health. According to the severity of pathogenicity, HPV is classified into low-risk and high-risk types, where low-risk types can lead to anal and genital warts, while persistent infection with high-risk types can cause precancerous lesions that eventually transform into tumors, such as cervical, penile, and oropharyngeal cancers, et al 12. HPV is the most common sexually transmitted infection, and the likelihood of HPV infection during the sexually active phase of the life of a man and a woman is 90% and 80%, respectively 3. Various evidences suggest that HPV infection is more common in the male population, and associated diseases caused by HPV infection are at a high prevalence among men of all ages around the world 46.

The burden of HPV-associated disease is huge, with anogenital warts incidence of 103∼168/100,000 per year in the male population and increasing rapidly at a rate of 2% per year 78, and the risk of HPV-associated anogenital disease is higher, particularly in the MSM population 9. Global Cancer Statistics 2020 reported age-standardized incidence rates of oropharyngeal and anal cancers in men of 1.8/100,000 and 0.5/100,000, respectively, and an estimated 22,000 additional cases of penile cancers per year globally, with approximately 40% of penile cancers being caused by HPV infection 1011.

Fortunately, HPV vaccines are effective in preventing infection among several oncogenic HPV subtypes and have become the most effective intervention for controlling HPV-related diseases 12. Studies have shown that HPV vaccination in girls and women’s populations has demonstrated protection against HPV-associated disease 13, and extending vaccination to boys and adult men may significantly reduce the burden of disease. The current quadrivalent and nine-valent HPV vaccines were licensed in the European Union in 2006 and 2015, respectively, as an important preventive measure against HPV-related diseases in men 14. The quadrivalent HPV vaccine was found to be consistently effective in preventing anogenital warts for at least 10 years, with an efficacy rate of 90% 15. A 10-year randomized controlled study in people aged 16∼26 years showed that in a heterosexual male population, the incidence of penile cancer, anal intraepithelial neoplasia, and anal cancer associated with HPV-6, HPV-11, HPV-16, and HPV-18 infections in HPV-vaccinated individuals was 43.4%, 59.0%, and 7.7% lower, respectively, compared to unvaccinated individuals 16. A meta-analysis that included 6,000 HPV vaccine recipients showed that HPV vaccination resulted in a 48% decrease in the detection of anal-genital warts in boys aged 15∼19 years and a 32% decrease in men aged 20∼24 years 17. Nonetheless, HPV vaccination rates remain very low in the male population. As of 2019, only 4% of men worldwide have been vaccinated against HPV 18. Due to the high cost of HPV vaccines, only a few countries around the world have included HPV vaccines for men and women in their immunization programs, such as Australia and the United States, and most countries, including China, have mainly focused on implementing vaccination in the girl and woman populations 19.

According to the Global Vaccination Agenda 2030, “making vaccines available to everyone, everywhere” and identifying “coverage and equity” of vaccination as a priority strategy 20. Gezimu W et al 21 concluded that vaccination of the female population alone is unlikely to alleviate human HPV infection and that males are still capable of carrying and transmitting the virus. In addition, the exclusion of men from vaccination program would also seem to run counter to gender equality initiatives and their sexual and reproductive health rights. The overall global goal is to eliminate HPV-related diseases by increasing HPV vaccination coverage and improving knowledge and awareness of HPV vaccination 18. Aggressive promotion of HPV vaccination in the male population could contribute to the global improvement of HPV-related diseases. Lack of HPV vaccine-related knowledge among men is a barrier to vaccination 22. Healthcare workers play a very important role in HPV vaccine awareness and vaccination and promotion due to their occupation 2324. Therefore, assess HPV vaccine awareness and willingness to vaccinate among male healthcare workers is important in the global control of HPV infection and elimination of cervical cancer.

In 2019, an international vaccine manufacturer completed a phase III clinical trial of HPV vaccination in men 25. In 2023, several HPV vaccine manufacturers in China are conducting phase III clinical trials of the male nine-valent HPV vaccine, and some have already opened blind, and soon the male HPV vaccine will be available in China **Error! Reference source not found.**27.

Therefore, this study aimed to assess the awareness and willingness of male healthcare workers to receive HPV vaccine and its influencing factors in Guilin City, a minority-populated area in southern China, to provide a reference basis for the government to formulate a promotion strategy after the male HPV vaccine is approved.

## 2. Materials and Methods

### 2.1. Study Design

A cross-sectional survey was conducted on male healthcare workers in medical institutions and Centers for Disease Control and Prevention (CDC) under the jurisdiction from September to October 2023 in Guilin city.

Guilin is located in the south of China, 1,960 kilometers from Beijing (the capital of China) and 380 kilometers from Nanning (the regional capital of Guangxi Zhuang Autonomous Region), with 17 counties/districts, 147 townships/streets, and 1,894 community/village committees, and a resident population of 4,950,000 people in 2023, with a concentration of more than 10 ethnic minorities, including Zhuang, Miao, Yao, Dong, and Hui et al.

### 2.2. Questionnaire Design and Sample Size Determination

This study consulted the literature related to HPV vaccination and willingness to vaccinate based on previous studies 2829, referred to the Chinese Expert Consensus on the Clinical Application of Human Papilloma virus Vaccine 30, prepared a self-administered web questionnaire, and revised it after consulting 12 epidemiologists. Convenience sampling method was used to select male healthcare workers from medical institutions and CDC under the jurisdiction of the survey area as survey respondents. Inclusion criteria: 1. age ≥ 18 years old; 2. male; 3. place of employment was medical institutions and CDC under the jurisdiction of Guilin City. Exclusion criteria: those who could not satisfy the above 3 conditions at the same time.

The sample size was calculated using the single population proportion formula: n=*μ*_α_^2^*P*(1-*P*)/*δ*^2^, where n = the desired sample size, and *P* = 77.90% (a result of survey of HPV vaccine-related knowledge in southern China by Chen S et al 31), *μ*_α_ = 1.96 (critical value at 95% confidence level), δ = 5% (the margin of error), n = (1.96)^2^ * 0.779 * (1-0.779) / (0.05)^2^ = 265. For possible substandard response during the study, the final sample size was increased by 10% to n=265 * (1+10%). by adding that, the total sample size was 292.

### 2.3. Survey content and methodology

The questionnaire consisted of four sections: The first part is an introduction to the purpose and significance of this study, the principles of informed consent and confidentiality. The second part contains the demographic information of the respondents, including age, area of residence, Institution level, Institution type, occupation, department type, education level, position, title, and per capita monthly household income. The third section covers the issue of HPV vaccine awareness. A total of eight questions were included, with a total of 15 scoring points, with 1 score for a correct choice and 0 score for a wrong choice, and respondents with a cumulative score of ≥ 9 scores were classified as being aware of the HPV vaccine. The fourth part assessed respondents’ attitudes toward receiving HPV vaccine, including willingness to vaccinate, willingness to pay for the vaccine, reasons that prevented vaccination, and factors that prompted vaccination. The questionnaire was distributed to medical institutions and CDC through the Questionnaire Star platform (a Chinese website specializing in online questionnaires). The survey started on 1 September and ended on 31 September. After reading the first part of the questionnaire to ensure informed consent, the respondents voluntarily completed the subsequent parts of the questionnaire. There were no incentives to complete the questionnaire, and all data collected were kept and analyzed confidentially.

### 2.4. Data Statistics and Analysis

Data were collated from the eligible questionnaires using the Excel 2021 software package and exported to the SPSS 26.0 software package for statistical analysis. Data collation is carried out by professionally trained personnel. Questionnaires that were not from the surveyed area, incomplete, had logical problems and took <120 seconds to complete were categorized as ineligible questionnaires and were given exclusion from this study. Quantitative information was described using mean ± standard deviation; disaggregated data were described using frequencies and percentages [n(%)]. HPV vaccination rate(number of people who have received HPV vaccine / total number of people surveyed * 100%). Frequency and percentage were used to describe the data, and the chi-square test or Fisher’ s exact test was used for categorical data. Multivariate analysis adopted stepwise logistic regression analysis. All variables with *p* ≤ 0.2 in single-factor analysis were included in the model, and the odds ratio (*OR*) and 95% confidence interval (*CI*) were calculated. Statistical significance was considered at *p* < 0.05. The significance level for entry (Slentry, SLE) was 0.05, and the significance level for stay (Slstay, SLS) was 0.10.

## 3. Results

### 3.1. Demographic Characteristics of Respondents

A total of 1,577 questionnaires were received, of which 1,430 were qualified, a qualification rate of 90.68%. Among the 1,430 respondents, The mean age was 37.80 ± 10.34(SD) years old, ranging from 18∼65 years old. Out of the 1,430 respondents, the majority, 626(43.78%) of the respondents were in the age category 25∼29 years; and more than half, 813 (56.85%) of them resided in the county and town; nearly two-thirds, 953 (66.64%) of the respondents worked in county or lower institutions; the majority of respondents were from level III and below level II medical institutions, accounting for 32.52% (465) and 30.84%(441), respectively; nearly two-thirds, 919 (64.27%) of respondents were doctors by occupation; more than half, 788 (55.10%) of respondents were in clinical departments; the vast majority 1,204 (84.20%) of respondents had a college/bachelor’s degree; and more than two-thirds, 1,024 (71.61%) of respondents had no position; more than half, (764) 53.43% of the respondents have junior titles or lower; and the majority, 580 (40.56%) of the respondents have a per capita monthly household income of between CNY 3,000 and 4,000(**Table 1**).

**Table 1.**
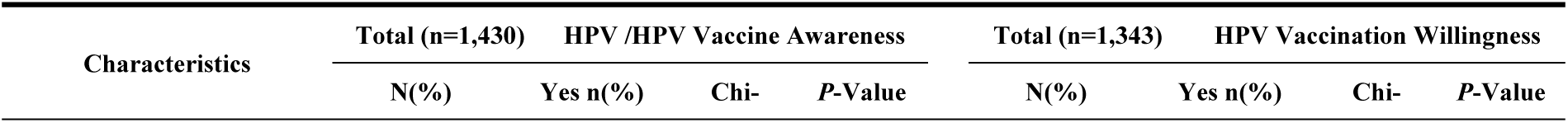
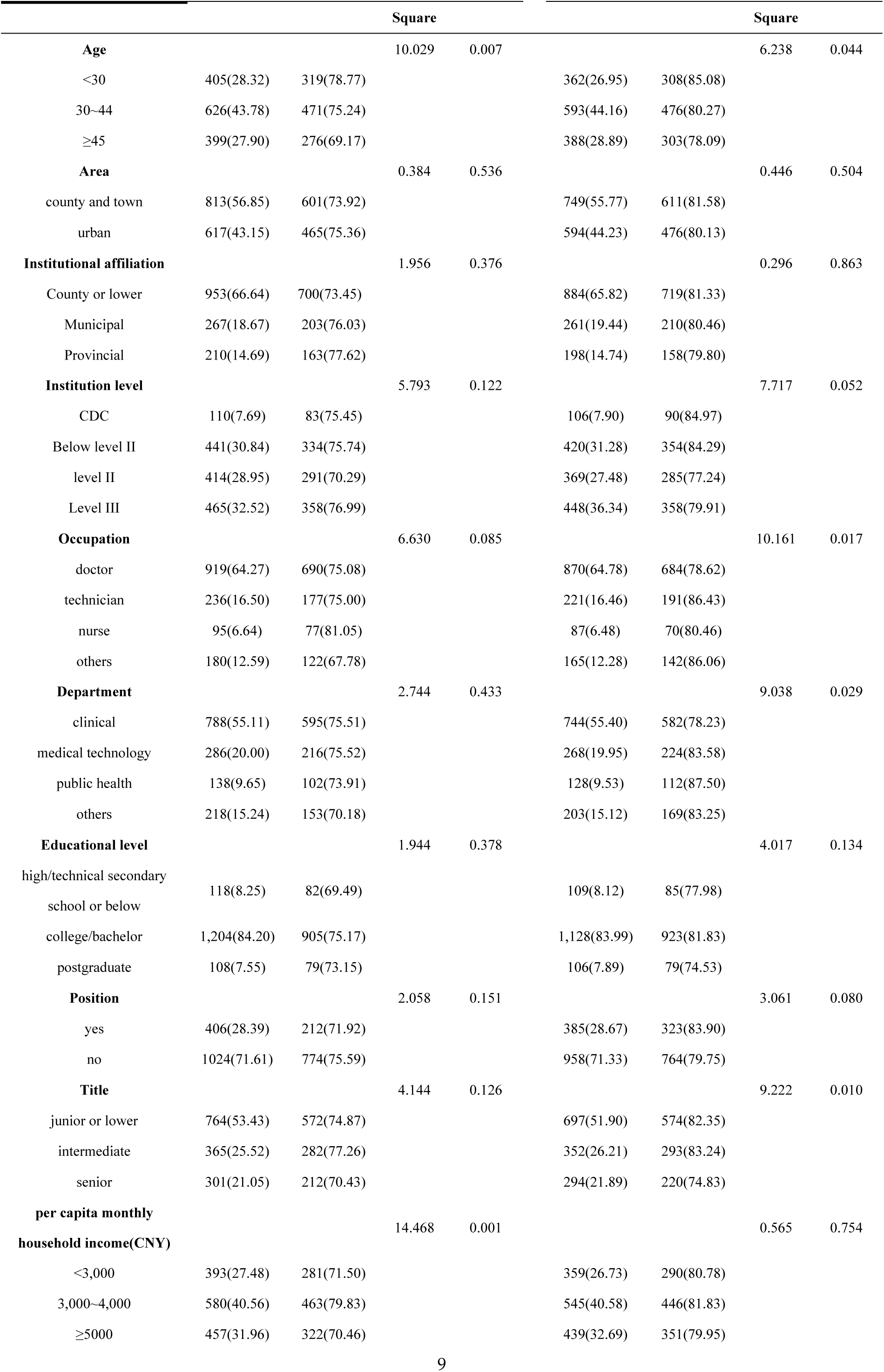
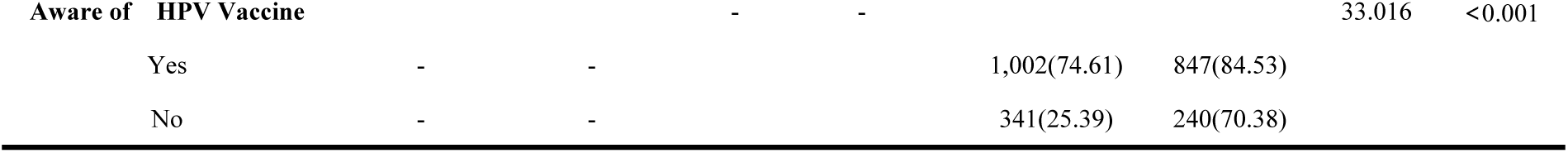
Demographic characteristics of the male healthcare workers, and the relationship between demographic characteristics and HPV vaccine awareness and HPV vaccination willingness.

### 3.2 respondents’ knowledge of HPV vaccine

The mean score of respondents’ HPV vaccine awareness was 9.47 ± 2.18. More than three-quarters, 1,066 (75.55%) of the respondents (scored ≥ 9) were categorized as being aware of HPV vaccine (**Fig 1**). Nearly all (98.11%, 1,403/1,430) of the respondents have heard of HPV. When HPV was mentioned, more than half, 725 (51.67%) and 809 (57.66%) of the respondents thought of STDs and genital warts, respectively, and more than four-fifths, 1,128 (80.40%) of the respondents thought of cervical cancer. Nearly two-thirds, 880 (62.72%) of the respondents believed that HPV infection would pose a high risk to their health. More than half, 797 (56.81%) of the respondents knew that HPV can be transmitted through direct or indirect contact, and a very small proportion (1.21%, 17/1,403) still believed that HPV was only transmitted through indirect contact. Less than one-fifth, 255 (18.18%) of the respondents knew the prevalence of HPV infection in the population. More than two thirds, 971(69.21%) of the respondents believed that they would experience discomfort after being infected with HPV. More than four-fifths, 1,233 (87.88%) of the respondents believed men can also getHPV. All 1,430(100%) of the respondents had heard of the HPV vaccine (**Table 2**).

**Fig 1.**
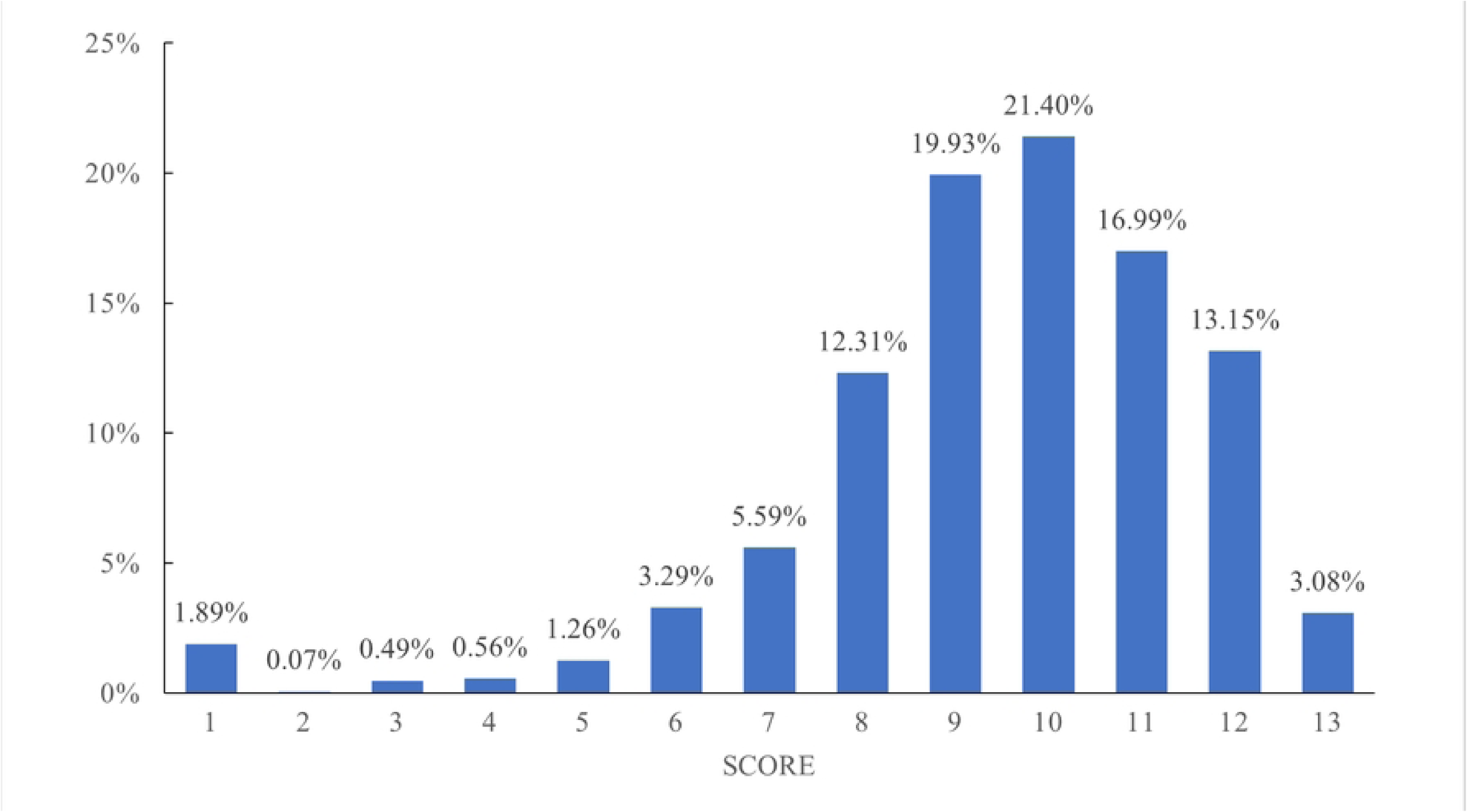
The score of HPV vaccine awareness among the male healthcare workers in the survey area of southern China, 2023 (n=1,430).

**Table 2.**
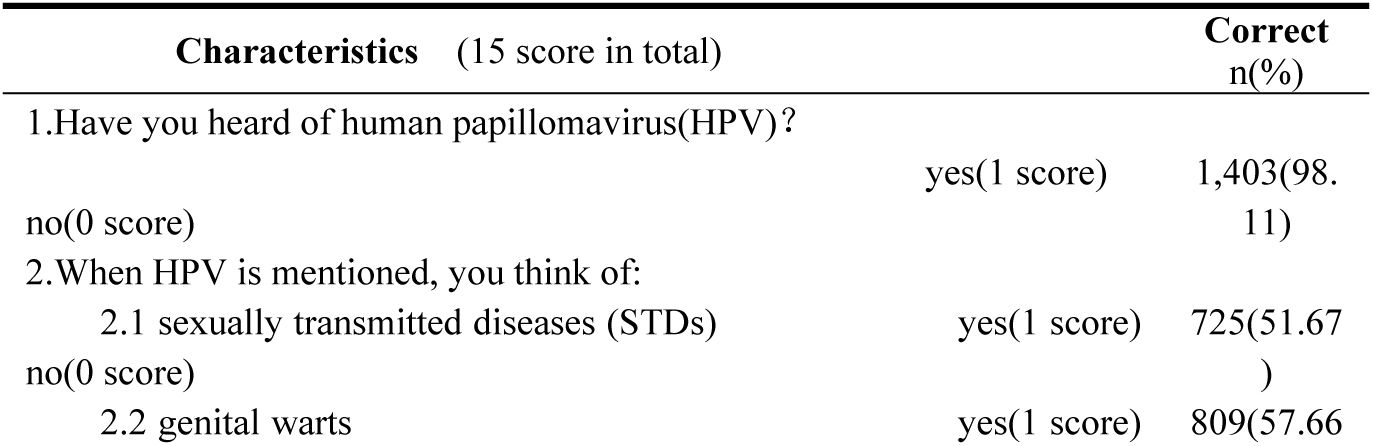
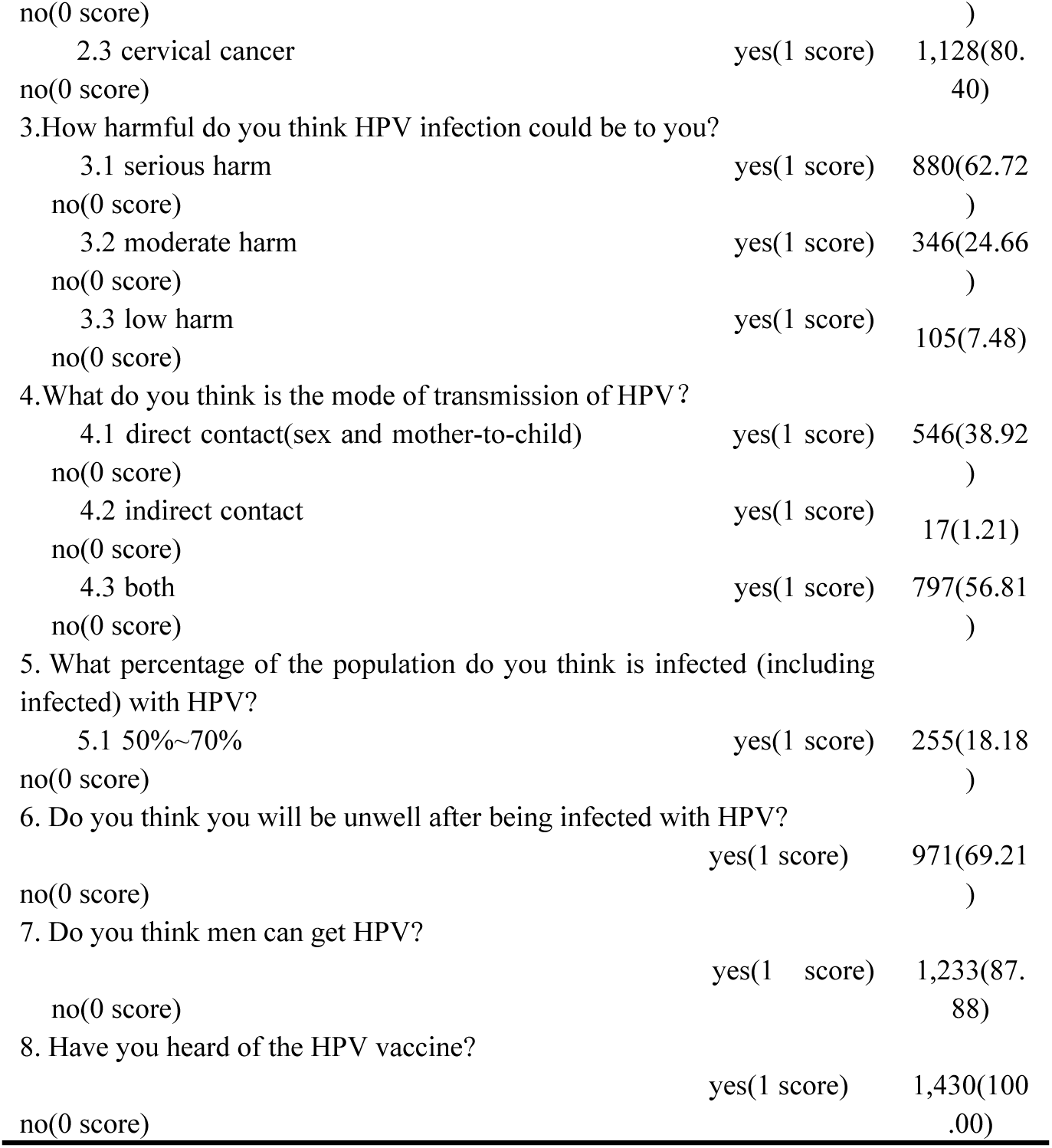
Knowledge-related questionnaire of the male healthcare workers on awareness associated factors of human papillomavirus vaccine among the respondents in the survey area of southern China, 2023 (n=1,430).

### 3.3 respondents’ attitudes towards receiving HPV vaccine

A very small proportion(6.08%, 87/1,430) of the respondents had received the HPV vaccine. Of the respondents who had not been vaccinated against HPV(n=1,343), three-quarters, 1,087(80.94%) of them were willing to be vaccinated against HPV. respondents’ willingness to be vaccinated against HPV was mainly due to the belief that the HPV vaccine could prevent HPV infection (91.44%, 994/1,087) and prevent genital warts (56.03%, 609/1,087) (**Fig 2**). Nearly three-fifths, 689(58.69%) of the respondents believed that the cost of HPV vaccines should be borne by the State, units of employment and individuals (**Fig 3**a), and nearly half 340(49.35%) believed that the proportion borne by the individual should be ≤ 20% (**Fig 3**b). More than half of the respondents who were willing to receive the (unvaccinated) HPV vaccine believed that the individual payment should be < CNY 200 (50.78%, 552/1,087) (**Fig 3**c). More than two-thirds 815(69.42%) of the respondents believed they can still be infected after HPV vaccination (**Fig 3**d). The main reasons why respondents were reluctant to receive the HPV vaccine were that they did not think they were at risk of HPV infection (133/256, 51.95%) and that the vaccine was expensive (80/256, 31.25%) (**Fig 4**). In addition, among the 627 respondents with daughters, the main measures that motivated respondents to get their daughters vaccinated against HPV were providing information about the purpose, importance, and effectiveness of the vaccine (578/627, 92.19%) and providing information about the safety of the vaccine (79.43%, 498/627)(**Fig 5**).

**Fig 2.**
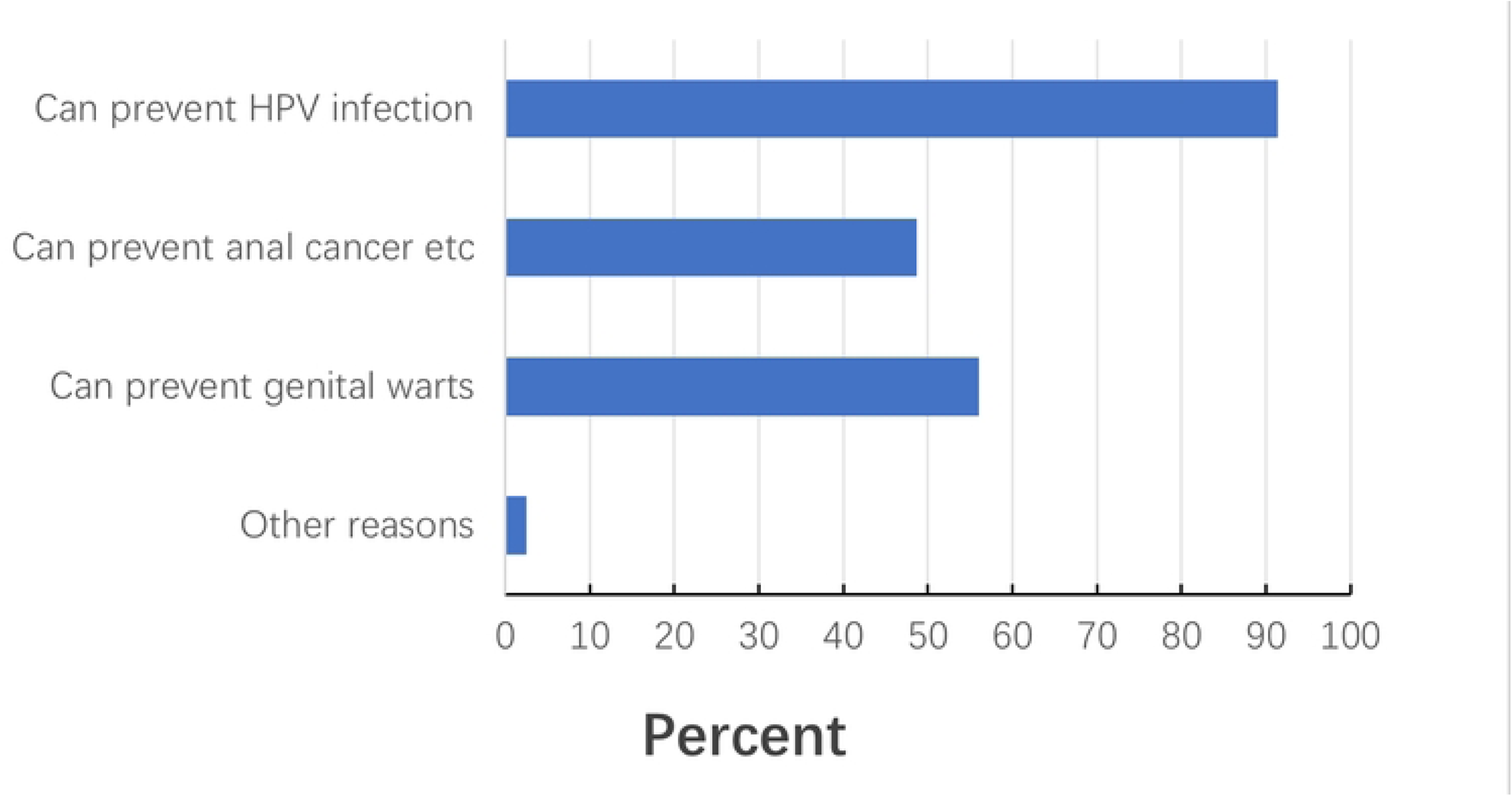
Reasons for male healthcare workers’ willingness to receive the HPV vaccine among the respondents in the Survey area of southern China, 2023 (n=1,087).

**Fig 3.**
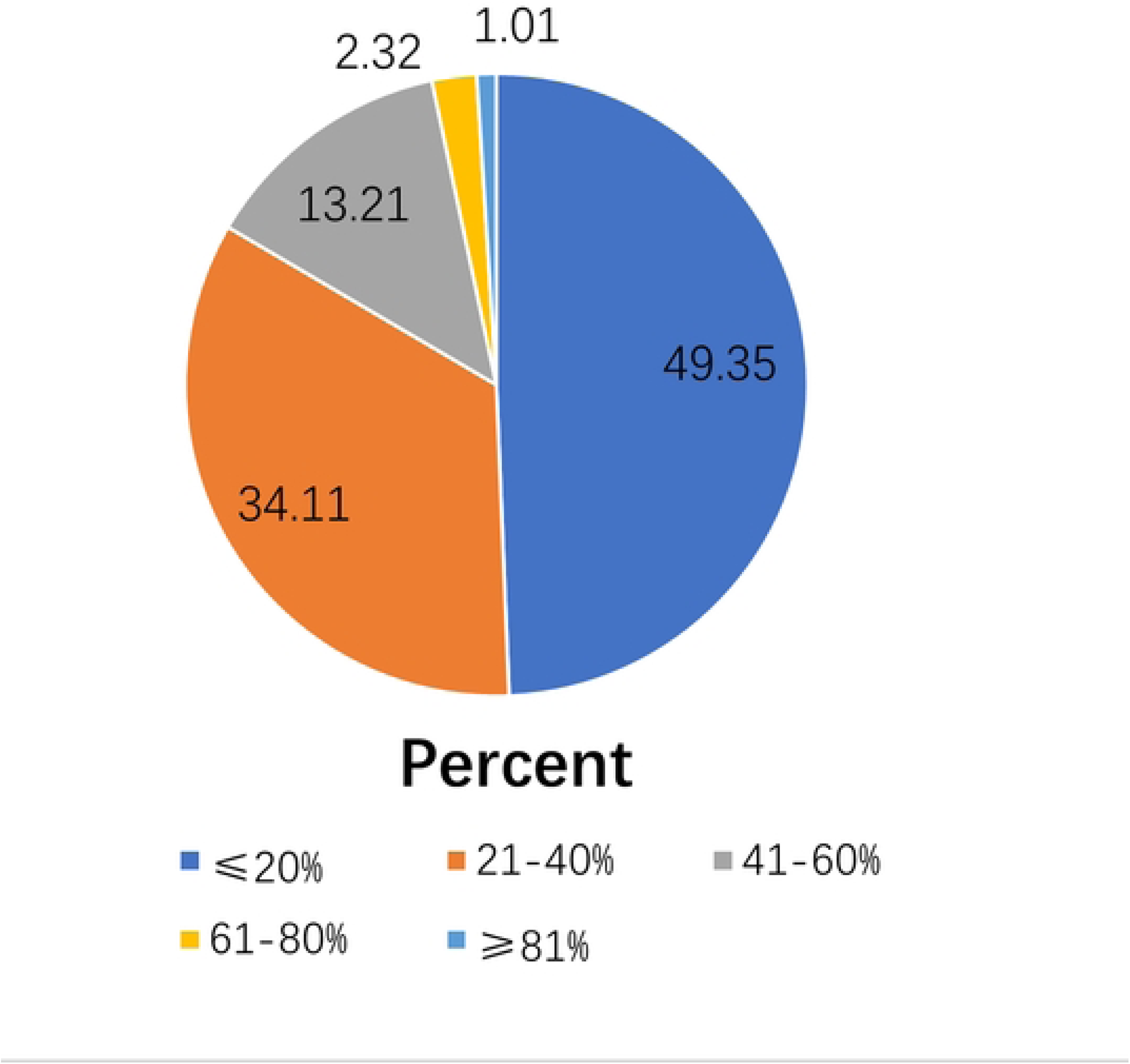
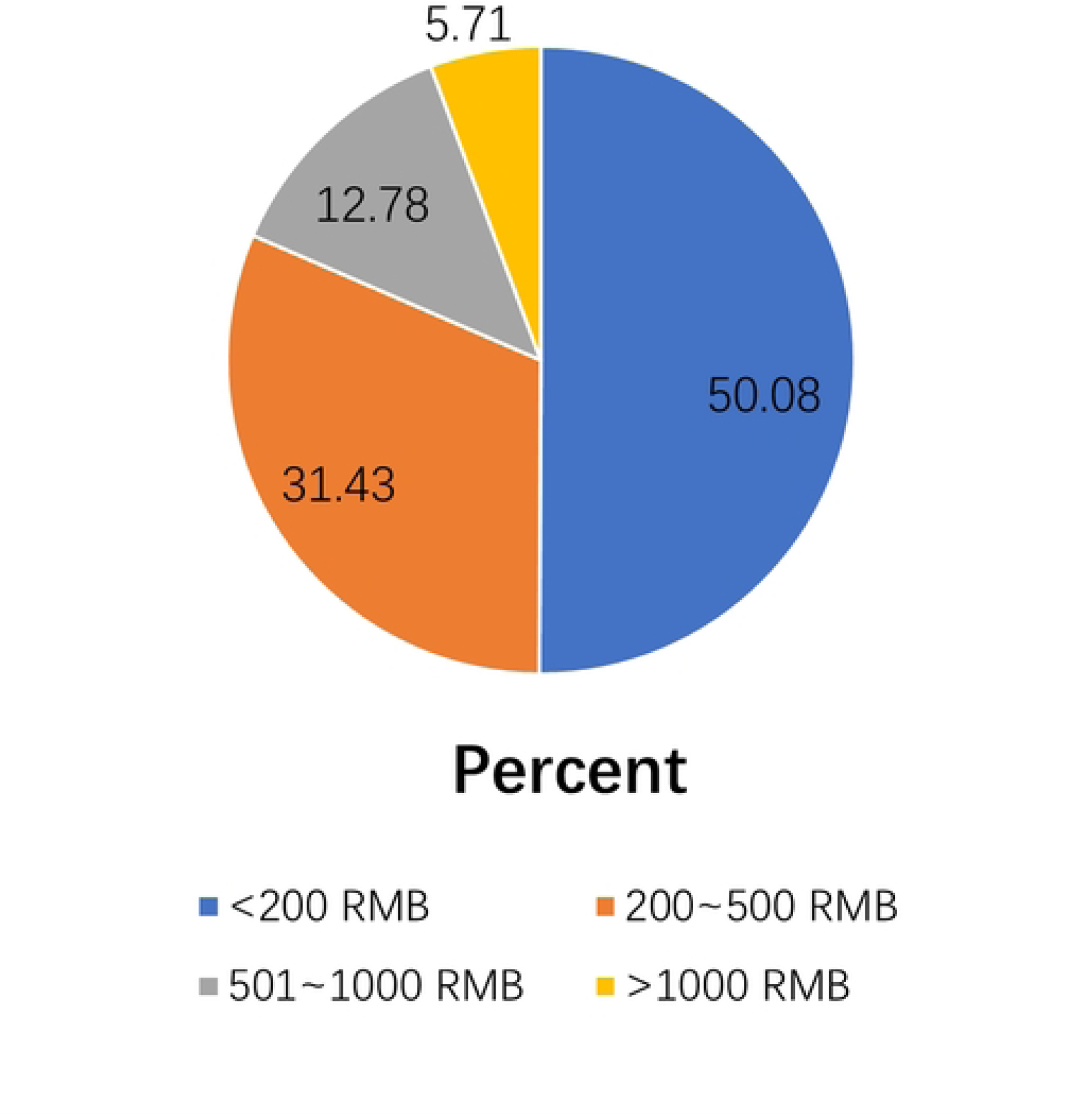
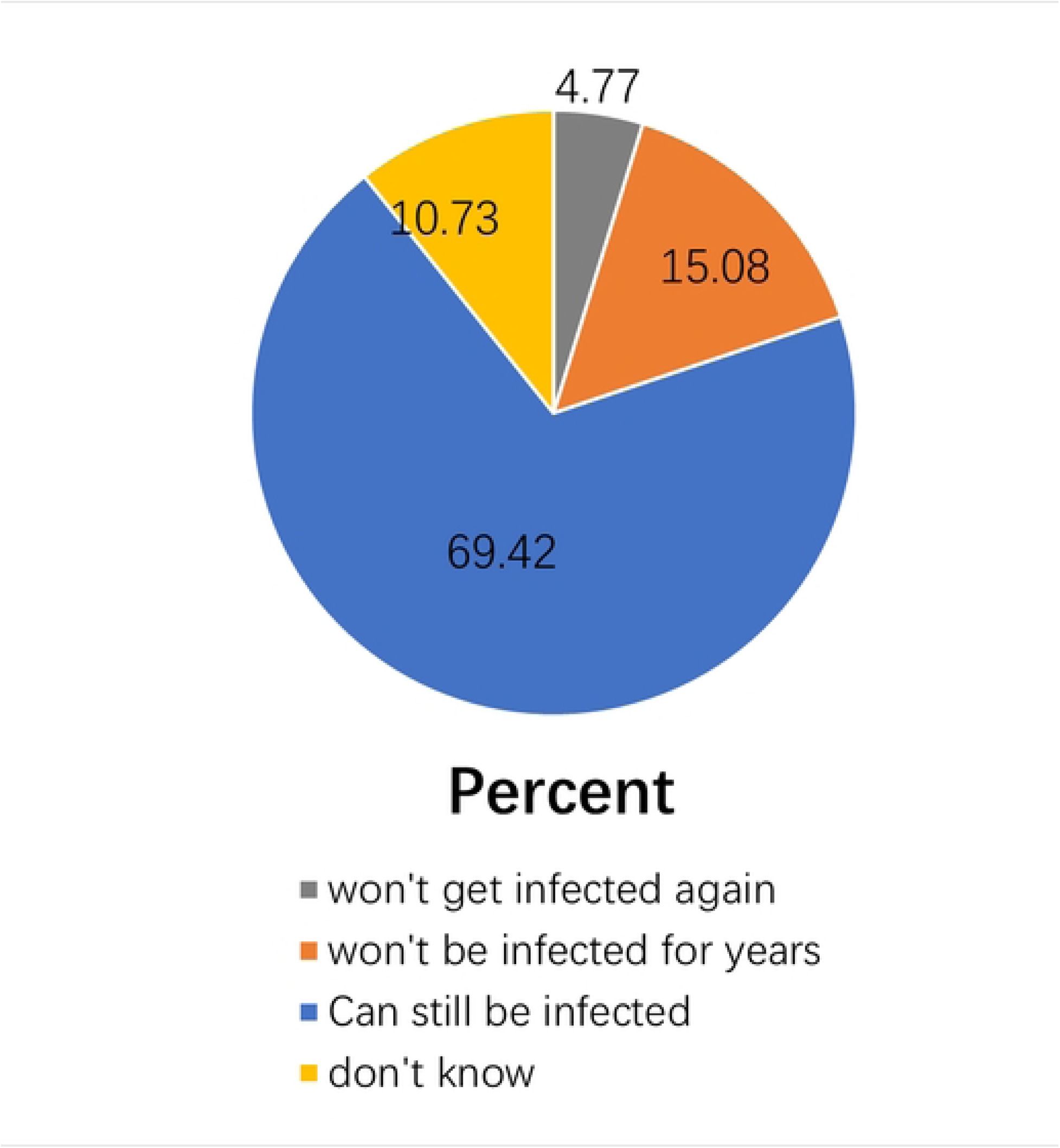
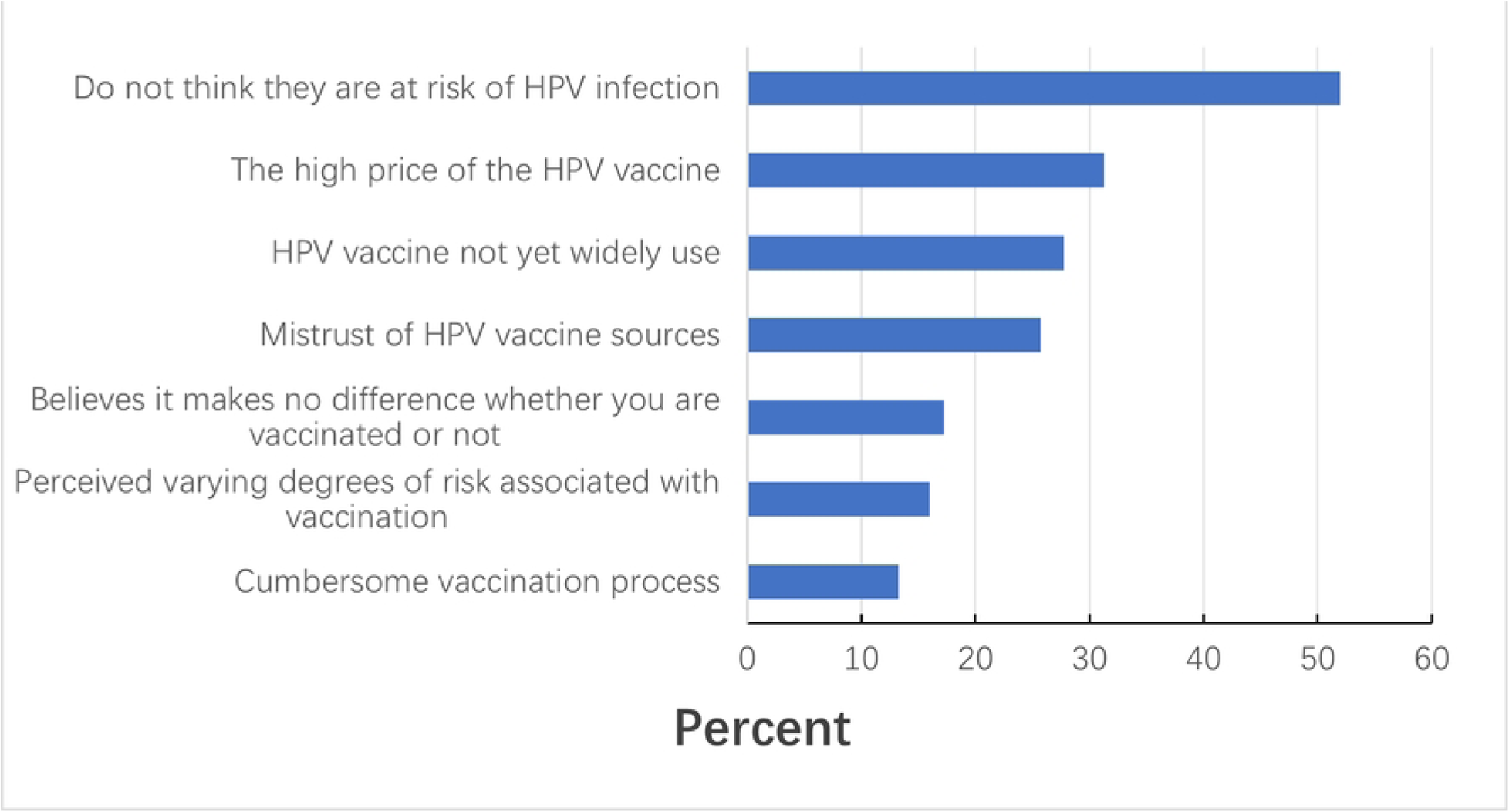
Behavior-related questionnaire of male healthcare workers on awareness, willingness, and associated factors of human papillomavirus vaccine in the survey area, 2023. (a) When asked, “How do you think who should bear the cost of HPV vaccination?”(n=1,174); (b) When asked, “What percentage do you think is appropriate for the individual to bear?”(n=689). (c) When asked, “How much are you willing to pay to have yourself vaccinated against HPV?” (n=1,087, excluding respondents had been vaccinated) (d) When asked, “Do you think you can still get HPV after getting the HPV vaccine?”(n=1,174).

**Fig 4.**
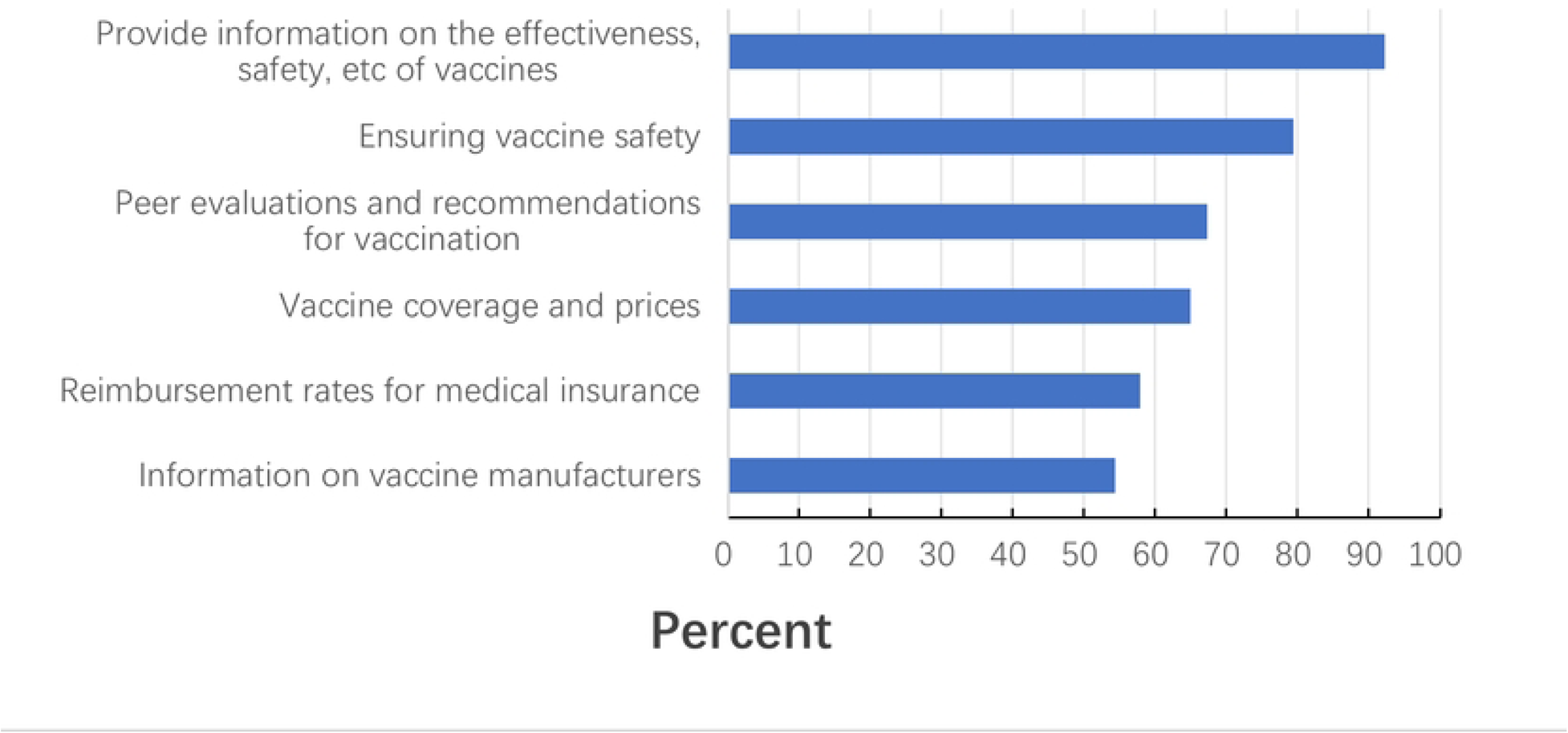
Reasons for male healthcare workers’ reluctance to receive HPV vaccine among male health workers in the survey area, 2023(n=256).

**Fig 5.**
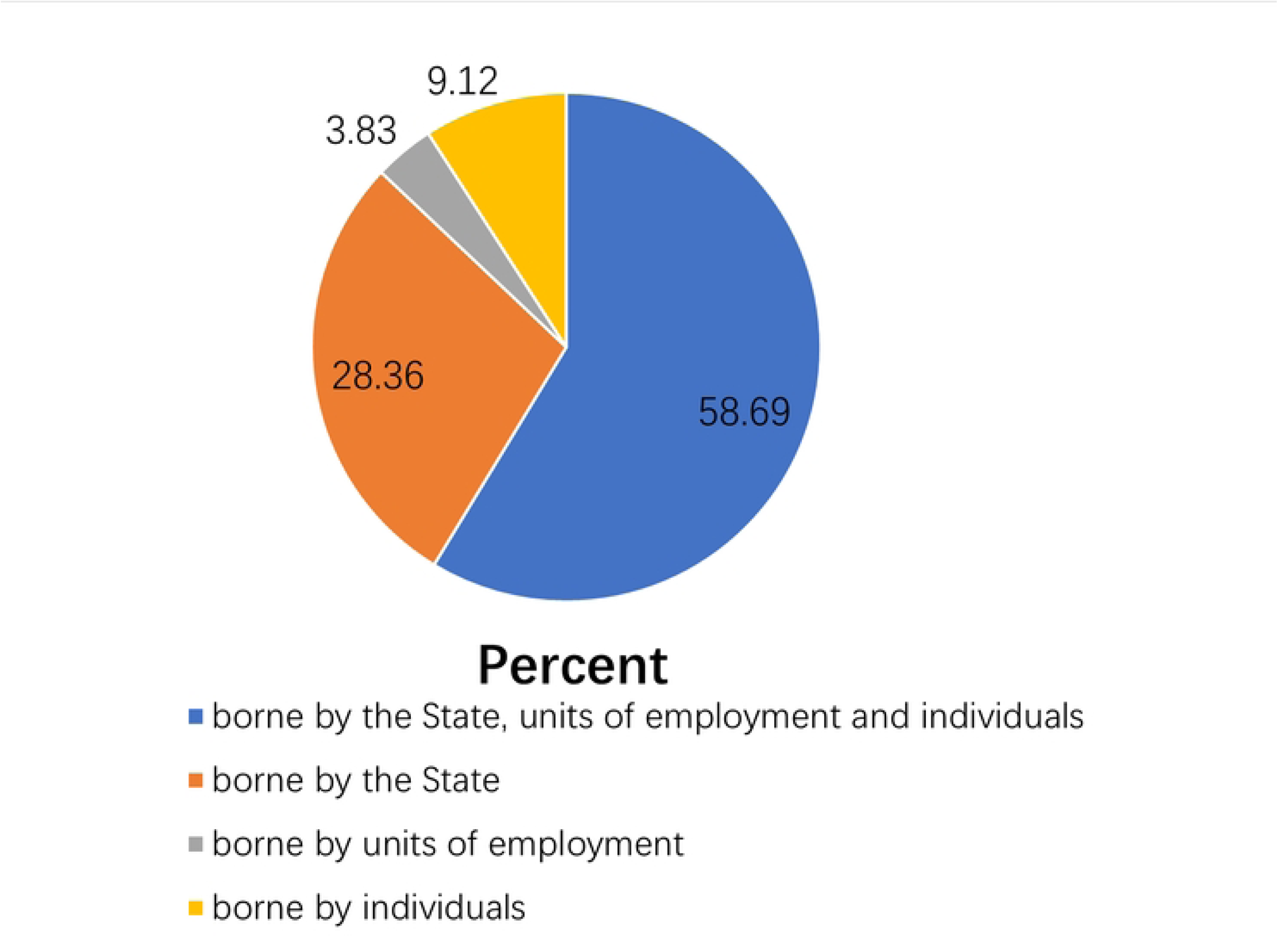
Measures that would promote male health workers to take their daughters for HPV vaccination among the male healthcare workers in the survey area, 2023. (n=627, respondents with daughters).

### 3.4 Multivariate Analysis of Factors Associated with HPV vaccine Awareness and HPV Vaccination Willingness

In multivariate analysis, HPV vaccine awareness was used as the dependent variable (0 = unaware, 1 = aware), and variables with *p* ≤ 0.2 in the single-factor analysis were included in the model, including age, institution level, occupation, position, title, and per capita monthly household income (**Table 1**). The results of the study showed that people aged 30∼44 and ≥ 45 years in the male health workers population have a low level awareness compared to < 30 (*OR* = 0.694, 95% *CI* = 0.495-0.972; *OR* = 0.477, 95% *CI* = 0.315-0.721). While, people whose with intermediate titles than those with junior titles or below, and with a per capita monthly household income of 3,000-4,000 CNY compared to < CNY 3,000 have a high awareness (*OR* = 1.530, 95% *CI* = 1.082-2.165; *OR* = 1.625, 95% *CI* = 1.191-2.217)(**Table 3**). And HPV vaccination willingness was used as the dependent variable (0 = unwilling, 1 = willing), and variables with *p* ≤ 0.2 in the single-factor analysis were included in the model, including age, institution level, occupation, department, educational level, position, title, and aware of HPV Vaccine (**Table 1**). The result of the study showed that people of technicians and others than doctors(*OR* = 1.822, 95% *CI* = 1.185-2.802; *OR* = 1.647, 95% *CI* = 1.009-2.688), those with a position than none (*OR* = 1.763, 95% CI = 1.242-2.503), and those aware of HPV vaccine than not (*OR* = 2.371, 95% *CI* = 1.767-3.182) in the male heath workers population were more willing to be vaccinated. While people whose with a senior title were less willing to be vaccinated than junior or below(*OR* = 0.579, 95% *CI* = 0.398-0.842) (**Table 4**).

**Table 3.** Multivariable analyses of factors associated with the awareness of HPV vaccine among the male health workers in the survey area of southern China, 2023 (n =1,430).

| Characteristics | Group | OR | 95%CI | P-Value |
| --- | --- | --- | --- | --- |
| Age | <30 | 1 |  |  |
|  | 30~44 | 0.694 | 0.495-0.972 | 0.034 |
| | $\geq 45$ | 0.477 | 0.315-0.721 | <0.001 |
| Title | junior or below | 1 |  |  |
|  | intermediate | 1.530 | 1.082-2.165 | 0.016 |
|  | senior | 1.257 | 0.847-1.867 | 0.256 |
| per capita monthly household income(CNY) | <3,000 | 1 |  |  |
|  | 3,000~4,000 | 1.625 | 1.191-2.217 | 0.002 |
| | $\geq 5,000$ | 0.961 | 0.677-1.364 | 0.824 |

**Table 4.** Multivariable analyses of factors associated with the willingness of HPV vaccine among the male health workers in the survey area of southern China, 2023 (n =1,343).

| Characteristics | Group | OR | 95%CI | P-Value |
| --- | --- | --- | --- | --- |
| Occupation | doctor | 1 |  |  |
|  | technician | 1.822 | 1.185-2.802 | 0.006 |
|  | nurse | 1.057 | 0.593-1.887 | 0.850 |
|  | others | 1.647 | 1.009-2.688 | 0.046 |
| **Position** | Yes | 1 |  |  |
|  | No | 1.763 | 1.242-2.503 | 0.002 |
| **Title** | junior or | 1 |  |  |
|  | intermediate | 1.021 | 0.715-1.458 | 0.911 |
|  | high | 0.579 | 0.398-0.842 | 0.004 |
| **Aware of HPV** | No | 1 |  |  |
|  | Yes | 2.371 | 1.767-3.182 | <0.001 |

## 4. Discussion

This study was conducted on male healthcare workers in ethnic minority-populated areas in southern China, aiming to understand the knowledge and attitudes of male healthcare workers in the surveyed areas towards HPV vaccination and the acceptance of HPV vaccine, and to grasp the awareness and acceptance of HPV vaccine and its associated factors of the survey respondents in the surveyed areas, so as to provide a basis for formulating promotion strategies for male HPV vaccines after its approved in mainland China.

The current survey showed that nearly three-quarters (74.55%) of the respondents in the survey area in 2023 are aware of the HPV vaccine, which is higher than the results of the survey conducted by Horio F et al 32 in Japan, and the results of the survey of a population of male university students in mainland China reported by Ran H et al 33, and the results of the survey conducted by Gol I et al and Sağtaş F et al 3435. The survey also revealed that almost all (98.11%) respondents answered “Yes” When asked “Have you heard of human papillomavirus(HPV)?”, which was higher than the results of the survey conducted by Suhaila K et al 36 among health care students and professionals in Nepal. When asked, “Do you think men can get HPV?” more than four-fifths (87.88%) of the respondents answered “yes”. All respondents in this survey had heard of the HPV vaccine, which is higher than the results of a survey conducted in Japan 32, which may be related to the increased publicity and promotion of the HPV vaccine in recent years in China. Despite the high awareness of HPV vaccine among respondents in the survey area, only 6.08% (87/1,430) of respondents received HPV vaccine, lower than the 18.90% in Europe 37, which may be related to the fact that male HPV vaccine has not been approved in the mainland China yet.

The result of multivariable analyses showed that HPV vaccine awareness was lower in the 30∼44 and ≥ 45 age groups compared to < 30, which may be related to the Immunization procedures for HPV vaccination, and also to the fact that young people have more frequent access to the Internet and more opportunities to acquire HPV-related knowledge from the Internet. A survey showed that the Internet is the main way for Algerian students to gain knowledge about HPV 38. People with intermediate titles had a higher level of awareness than those with junior titles or below, which may be related to work experience. Awareness of HPV vaccine was higher among those with a per capita monthly household income of CNY 3,000∼4,000 than those with < CNY 3,000, which may be related to health awareness, which is consistent with the findings of Zhou W et al 39.

The survey also founded that although near to three-quarters, 815(69.42%) of the respondents (Including those willing to be vaccinated and those already vaccinated against HPV) believed that they could still be infected with HPV after HPV vaccination, the majority, 1,087(80.94%) of the unvaccinated respondents were still willing to receive HPV vaccine. The vaccination willingness is close to the survey results of the Northern Nigerian healthcare students (81.5%), and the finding of mainland China’s gay and lesbian community (85.1%)4041, which may be related to the respondents’ sense of the dangers of HPV and the importance of HPV vaccine. This study also showed that 62.72% (880/1,430) of the respondents believed that HPV caused serious bodily harm, as well as the main reason for their willingness to receive the HPV vaccine was the belief that the HPV vaccine can prevent HPV infection (91.48%, 1,074/1,174). In contrast, the main barriers to HPV vaccination for respondents were that they did not perceive themselves to be at risk of HPV infection (51.95%, 133/256) and that the vaccine was expensive (31.25%, 80/256), which is similar to the findings in Turkey 42 and Singapore 43.

The result of multivariable analyses also showed that technicians and other occupations had a higher willingness to vaccinate than doctors, while people with senior titles had a lower willingness to vaccinate than junior titles and below, which may be related to the fact that doctors and those with higher titles are more likely to be skeptical about vaccine safety and efficacy 44. And higher among people with position than no position, which may be related to that they are more likely to have higher levels of social responsibility and self-awareness of health management, which is need further research. People aware of HPV vaccine had a higher willingness to vaccinate than those unaware, is in line with the results of a survey in the Greek region 45, which suggests that increased awareness-raising and training about HPV vaccine could contribute to the increase in willingness to vaccinate.

The study also revealed that the most effective measures that would motivate people with daughters to get their daughters vaccinated against HPV were providing information about the purpose and effectiveness of the vaccine (92.19%), and the safety of the vaccine (79.43%), peer’ s evaluations and recommendations of the vaccine (67.30%), and the price of the vaccine (64.91%), which is similar to the findings of Mortensen GL et al46. On the one hand, it suggests that respondents focus on the effectiveness and safety of the HPV vaccine, and the other hand, it suggests that vaccine’ s price plays an important role in the decision maker’s final decision to vaccinate or not 47. In this survey, 58.69% of the respondents believed that the cost of HPV vaccine should be borne by the state, the units of employment and individuals, and 81.52% of the respondents were willing to pay < CNY 500 for HPV vaccination. According to statistics, among 107 countries that support HPV vaccination program, only 33 countries have implemented HPV vaccination program for both genders, while in mainland China, due to the large population size and the tight supply of vaccines, HPV vaccines are still voluntarily and self-funded, the price of the vaccine is one of the most important factors affecting vaccination, suggesting that setting reasonable HPV vaccination prices can help increase vaccination rates3947.

## 5. Conclusions

In summary, awareness of HPV vaccine and willingness to vaccinate among male healthcare workers in ethnic minority-populated areas in southern China are in high level, but the perception is not thorough enough. The belief that they are not at risk of HPV infection and the high price of the HPV vaccine are the main obstacles to receiving the HPV vaccine. Providing with information on HPV vaccine efficacy and safety, and lowering the price of HPV vaccine could promote HPV vaccination. Increasing awareness of HPV vaccines can increase willingness to vaccinate against HPV. It is suggested that HPV vaccine training should focus on older age groups, high title and low title and below, doctors and low-income populations in late time. The health education should focus on the effectiveness and safety of the vaccine. Government should consider incorporating the HPV vaccine into the immunization program or including the vaccine in the health insurance subsidy catalog to promote HPV vaccination after the approval of male HPV vaccine in mainland China.

## 6. Strengths and limitations

Strengths of this study are as follows: 1. Male HPV vaccine will be approved in China soon, and the study on awareness and willingness to be vaccinated in ethnic minority-populated areas in southern China will be of greater significance for the later promotion of HPV vaccine in southern China. 2. Healthcare workers play an important role in HPV prevention and control campaigns and later HPV vaccine promotion due to their special professional and academic reasons. 3. This survey is characterized by a wide coverage and a large sample size, and the results are highly representative. Since the study is cross-sectional, a causal relationship cannot be shown, and recollection and social desirability bias may potentially have an impact in the online questionnaire survey, which may affect the study results to a certain extent. In addition, the study is targeted at ethnic minority neighborhoods in small cities in southern China, and the generalization of the results is somewhat limited.

## Author Contributions

Chunlin Qin: Study design, Data curation, Formal analysis, Investigation, Writing original draft, Writing review & editing. Qingqing Liang: Study design, Conceptualization, Methodology, Writing review & editing, Coordination of administrative affairs. Nian Jiang: Data curation, Investigation, Writing original draft. Yun Zhou: Data curation, Investigation, Methodology, Writing review & editing. Guorong Tang: Study design review, Conceptualization, Formal analysis, Methodology, Writing review, Coordination of administrative affairs.

## Institutional Review Board Statement

The present study was conducted in accordance with the guidelines of the Declaration of Helsinki. The Ethics Committee of Guilin Centre for Disease Control and Prevention provided the necessary ethical approval, Ref No GLCDC-Medical Ethics-202306 with the date 26/07/23.

## Informed Consent Statement

Informed consent was obtained from all subjects involved in the study.

## Data Availability Statement

The datasets involved in the current study are not publicly available due to privacy but are available from the author Chunlin Qin on reasonable request.

## Acknowledgments

Thanks to all respondents and investigators for their help.

